# VOYAGER: an international consortium investigating the role of human papilloma virus and genetics in oral and oropharyngeal cancer risk and survival

**DOI:** 10.1101/2025.02.17.25322399

**Authors:** Mark Gormley, Ashim Adhikari, Tom Dudding, Miranda Pring, Katrina Hurley, Gary J Macfarlane, Pagona Lagiou, Areti Lagiou, Jerry Polesel, Antonio Agudo, Laia Alemany, Wolfgang Ahrens, Claire M Healy, David I Conway, Cristina Canova, Ivana Holcatova, Lorenzo Richiardi, Ariana Znaor, Andrew F Olshan, Rayjean J Hung, Geoffrey Liu, Scott V Bratman, Xiaobei Zhao, Jeremiah Holt, Ricardo Cortez, Valerie Gaborieau, James D McKay, Tim Waterboer, Paul Brennan, D. Neil Hayes, Brenda Diergaarde, Shama Virani

## Abstract

Head and neck cancer (HNC) is the sixth most common cancer globally. Incidence and survival rates vary significantly across geographic regions and HNC tumor subsites. This is partly due to differences in risk factor exposure, including tobacco smoking, alcohol consumption and human papillomavirus (HPV) infection, alongside detection and treatment strategies. The VOYAGER (human papillomaVirus, Oral and oropharYngeal cAncer GEnomic Research) consortium is a collaboration between five large North American and European studies which generated data on more than 10,000 participants (7,233 cases and 3,297 controls). The primary goal of the collaboration was to improve understanding of the role of HPV and genetic factors in oral cavity and oropharyngeal cancer risk and outcome. Demographic and clinical data collected by the five studies were harmonized, and HPV status was determined for the majority of cases. In addition, almost 1,000 tumors were sequenced to characterize somatic alterations. The resulting comprehensive biomedical resource can be utilized to answer critical outstanding research questions to help improve HNC prevention, early detection, treatment, and surveillance.

**Highlights:**

- VOYAGER is a large harmonized international resource for advancing head and neck cancer research.
- Comprehensive demographic, clinical and genetic data to support studies of head and neck cancer risk and survival.
- Standardized data harmonization to enable robust analyses across multiple populations.
- Long-term follow-up to facilitate the development of prognostic and predictive biomarkers.

## Introduction

### Head and Neck Cancer Genomic Epidemiology

Head and neck cancer (HNC) which is primarily squamous cell carcinoma, includes cancers of the oral cavity, pharynx and larynx [1, 2]. Globally, the incidence of oral and oropharyngeal cancer is estimated at 8.0 and 2.0 per 100,000, respectively, and is predicted to increase by 30% by 2030 [3, 4]. Five-year survival remains poor, averaging between 40 – 50% with hypopharynx cases experiencing the worst outcomes [5]. Incidence and survival rates vary significantly across geographic regions and tumor subsites, partly due to differences in risk factor exposure. Established HNC risk factors include tobacco smoking and alcohol intake, which together account for a similar population attributable risk for both oral (64%) and oropharyngeal (72%) cancer [6]. However, human papilloma virus (HPV) infection, particularly high-risk subtype 16, has emerged as another major risk factor for oropharyngeal cancer [7–9]. Worldwide, it is estimated that around 52,000 incident HNC cases are caused by a persistent HPV infection each year, with attributable fractions highest in high-income countries in North America and Europe [10–13].

Given the decline of tobacco use in developed countries, the incidence rate of HPV driven [HPV(+)] oropharyngeal cancer is now surpassing that of oral cancer [9–12, 14]. HPV(+) oropharyngeal tumors are considered distinct entities, demonstrating more favorable treatment response and prognosis compared to non-HPV related oropharyngeal cancer [HPV(-)] [8, 12, 15–17]. This is likely due to differences in etiology, and patient and tumor characteristics, with HPV(+) oropharyngeal tumors presenting more frequently in younger individuals (<65 years), and in those reporting higher numbers of sexual partners with reduced cumulative tobacco exposure compared to HPV(-) cases [12, 18]. However, only a small proportion of those with an oral HPV infection will develop HNC and despite better long-term survival, up to 25% of patients still develop disease recurrence within 5 years after initial diagnosis [19]. To improve prevention, early detection and prognosis, a better understanding of the role of host genetics and interactions with modifiable risk factors, such as tobacco and alcohol use in oral and oropharyngeal cancer risk and survival is required [20].

HPV driven carcinogenesis is characterized by increased expression of the viral oncogenes E6 and E7, leading to increased degradation of tumor suppressor proteins p53 and Rb, respectively and loss of cell cycle activation. This can result in genomic instability and resistance to apoptosis [21, 22]. HPV(+) and HPV(-) head and neck tumors harbor a similar burden of somatic variants. However, HPV(+) oropharyngeal tumors have been reported to carry fewer copy-number alterations, suggesting a higher degree of genomic stability [23–26]. Genome profiling studies have provided a list of genes that are recurrently mutated in HNC, including *TP53*, *CDKN2A* (which encodes for p16^INK4^), *NOTCH1* and *PIK3CA* [23, 24, 27–29]. Genes recurrently mutated in HPV(+) oropharyngeal cancer are related to epithelial structure and differentiation, in addition to *RB1* (encoding the Rb protein) [12, 23–26]. The presence or absence of particular somatic alterations in tumors may be good markers of cancer prognosis and response to treatment. Therefore, there is still a need to identify novel somatic driver alterations, particularly as relatively few HPV(+) oropharyngeal cancer cases have been sequenced to date [24, 30, 31]. Identification of molecular markers associated with prognosis could facilitate better clinical decision making, including the use of de-escalation treatment strategies among those at lower risk of recurrence or progression as a means to improve quality of life and, conversely, more aggressive treatment in those deemed at higher risk.

Compared to other major cancer sites such as breast, lung and colorectal, HNC is relatively rare, hampering research efforts. The collaboration of the five North American and European studies in the VOYAGER (human papillomaVirus, Oral and oropharYngeal cAncer GEnomic Research) consortium has resulted in a large comprehensive biomedical resource that can be utilized to help resolve outstanding research questions and improve HNC prevention, early detection, treatment, and surveillance.

## Methods

### The VOYAGER Consortium: Design

In 2016, the VOYAGER consortium was established, bringing together five large North American and European studies (**Figure 1**) with a focus on oral and oropharyngeal cancers. The primary goal of the collaboration was to improve understanding of the role of HPV and genomic factors in oral cavity and oropharyngeal cancer risk and outcome. The project was funded by the US National Institute of Dental and Craniofacial Research (NIDCR; R01DE025712).

**Figure 1.**
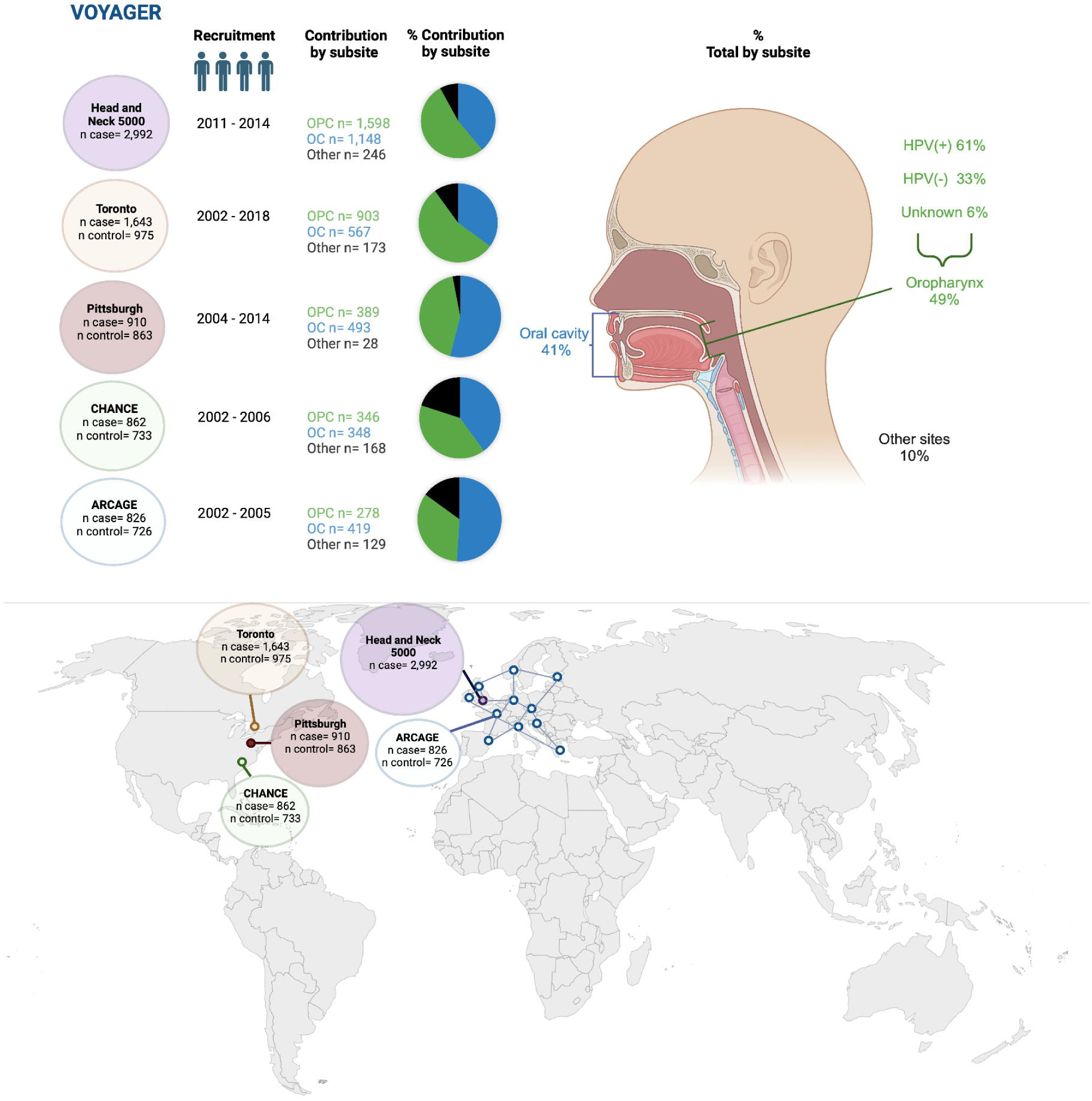
Overview of the five studies included in VOYAGER. Key: HPV, human papilloma virus; OPC, oropharyngeal cancer (green); OC, oral cancer (blue); Other sites (black). *Created in BioRender. Gormley, M. (2026)* https://BioRender.com/j07p735

The VOYAGER consortium includes 10,530 participants in total (6,489 oral and oropharyngeal cancer cases, 744 other head and neck cancer cases, and 3,297 controls), with detailed demographic, risk factor and clinical data. The five studies comprising VOYAGER have been previously described and are: (a) the Alcohol-related cancers and genetic susceptibility in Europe (ARCAGE) study [32], (b) the Toronto Mount Sinai Hospital-Princess Margaret (MSH-PMH) study (Toronto) [33], (c) the University of Pittsburgh case-control study on head and neck cancer (Pittsburgh) [34], (d) the Carolina Head and Neck Cancer Epidemiology (CHANCE) study [35], and (e) the Head and Neck 5000 study (HN5000) [36]. Ethical approval was obtained as described in the **Declarations** section below.

Each study contributed oral cavity and oropharyngeal cancer cases as defined by the following ICD-10 codes: oral cavity (C00.3-C00.6, C00.8-C00.9, C02.0-C02.3, C02.8, C02.9, C03.0-C03.9, C04.0-C04.9, C05.0, C05.8, C05.9, C06.0-C06.9), and oropharynx (C01-C01.9, C02.4, C05.1-C05.2, C09.0-C10.9). Other HNC subsites that are also part of the VOYAGER dataset include: hypopharynx (C12.0-C13.9; N=357), larynx (C32; N=107), unknown primary (C80; N=103), pharynx not otherwise specified (NOS) (C14.0; N=72), external lip (C00.0-00.2; N=26), overlapping cases (N=44), and “Other” sites (nasal cavity, N=1; nasopharynx N=4; other ear/ bone/ skin/ lacrimal duct and gland, N=4; overlapping lesion, N=3; paranasal sinuses, N=4; salivary glands, N=2, head and neck cancer NOS, N=13; and, carcinoma in situ, N=4). Demographic information, including age, sex, ethnicity, geographic region and education level, and information on established risk factors, e.g., smoking and alcohol consumption history, were shared. Clinical variables included ICD code, tumor, nodal and metastasis status, HPV status (defined by p16 or HPV DNA), vital status, follow-up time, and treatment information. Given that all cases were diagnosed between 2002 – 2018, the American Joint Committee on Cancer (AJCC) 7^th^ edition was used, mirroring the staging used in clinical practice during that time period [37].

### Data Generation and Harmonization

The VOYAGER consortium generated several different types of data including clinical, demographic and behavior variables. Data harmonization was conducted for variables that were centralized across all studies, as illustrated in **Tables 1** and **2**. Upon receipt of data, cleaning and validation checks were conducted to identify inconsistencies, outliers, and missing values. Follow-up data was carefully collated and harmonized to capture follow-up times, progression events, vital status and cause of death to facilitate high-quality prognostic research. A comprehensive data dictionary was developed to specify final definitions, formats, permissible values, coding schemes and classifications (i.e., for categorical variables). Version control was implemented to ensure consistency of data use across analyses.

**Table 1.**
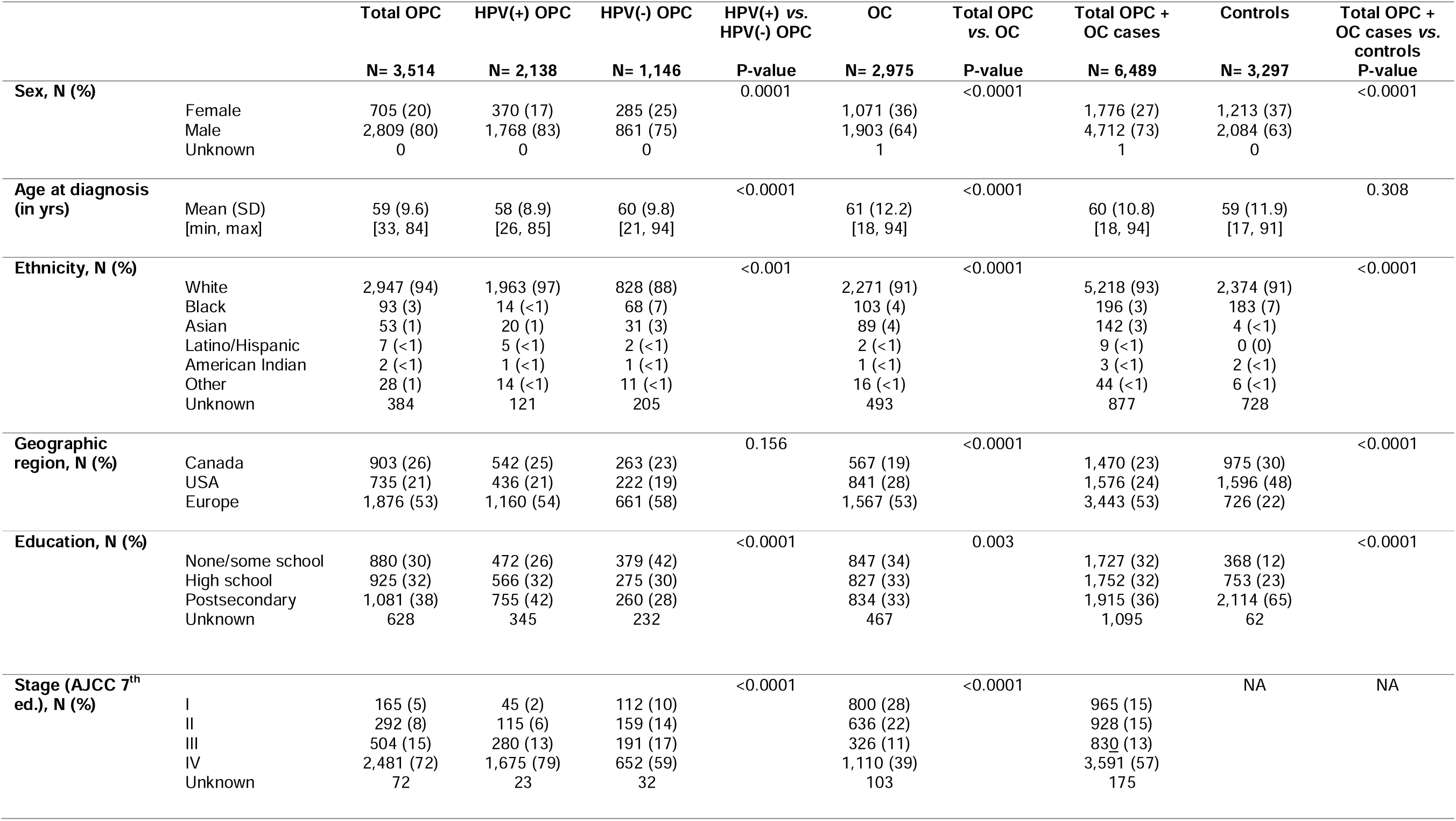
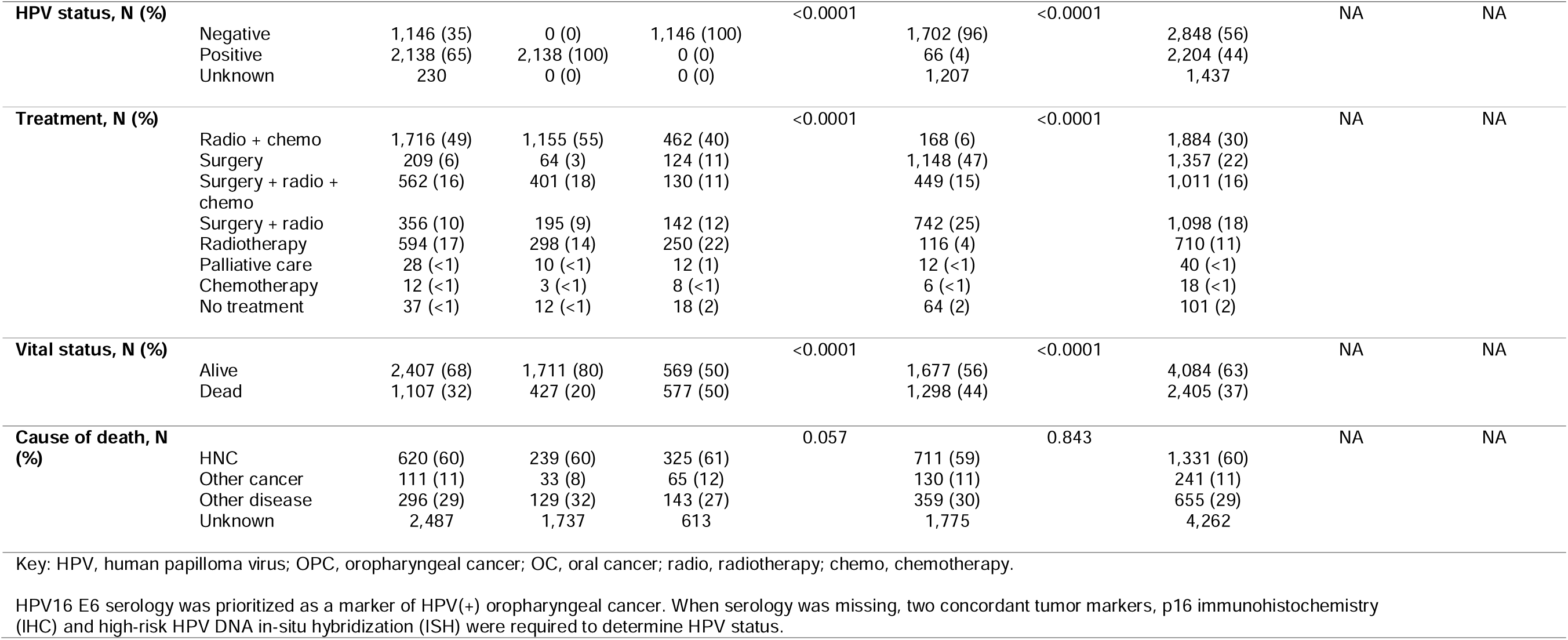
Clinical and demographic characteristics of the oral and oropharyngeal cancer cases and controls included in VOYAGER, stratified by subsite and HPV status.

**Table 2.** Information on established risk factors for the oral and oropharyngeal cancer cases and controls included in VOYAGER, stratified by subsite and HPV status.

|  |  | Total OPC<br>N= 3,514 | HPV(+) OPC<br>N= 2,138 | HPV(-) OPC<br>N= 1,146 | HPV(+) vs.<br>HPV(-) OPC<br>P-value | OC<br>N= 2,975 | Total OPC<br>vs. OC<br>P-value | Total OPC + OC<br>cases<br>N= 6,489 | Controls<br>N= 3,297 | Total OPC + OC<br>cases vs. controls<br>P-value |
| --- | --- | --- | --- | --- | --- | --- | --- | --- | --- | --- |
| **Smoking**<br>**status, N (%)** | Current | 862 (28) | 277 (15) | 498 (51) | <0.0001 | 988 (37) | <0.0001 | 1,850 (32) | 545 (16) | <0.0001 |
|  | Former | 1,425 (46) | 949 (51) | 381 (39) |  | 1,001 (38) |  | 2,426 (42) | 1,298 (40) |  |
|  | Never | 787 (26) | 638 (34) | 103 (10) |  | 671 (25) |  | 1,458 (26) | 1,437 (44) |  |
|  | Unknown | 440 | 274 | 164 |  | 315 |  | 755 | 17 |  |
| **Alcohol**<br>**drinking**<br>**status, N (%)** | Current | 1,984 (64) | 1,212 (65) | 643 (65) | <0.0001 | 1,618 (61) | <0.0001 | 3,602 (62) | 1,888 (58) | <0.0001 |
|  | Former | 463 (15) | 233 (12) | 180 (18) |  | 387 (14) |  | 850 (15) | 666 (20) |  |
|  | Never | 376 (12) | 230 (12) | 95 (10) |  | 437 (16) |  | 813 (14) | 722 (22) |  |
|  | Never/ Former * | 278 (9) | 211 (11) | 67 (7) |  | 228 (9) |  | 506 (9) | 0 (0) |  |
|  | Unknown | 413 | 252 | 161 |  | 305 |  | 718 | 21 |  |
| **Body mass**<br>**index, in**<br>**kg/m<sup>2</sup>, N (%)** | Healthy (18.5 to <25) | 1,034 (35) | 508 (28) | 452 (47) | <0.0001 | 1,057 (42) | <0.0001 | 2,091 (38) | 433 (27) | <0.0001 |
|  | Underweight (<18.5) | 91 (3) | 18 (<1) | 68 (7) |  | 96 (4) |  | 187 (3) | 12 (<1) |  |
|  | Overweight (25.0 to <30) | 1,080 (36) | 744 (40) | 280 (29) |  | 820 (33) |  | 1,900 (35) | 623 (39) |  |
|  | Obese (>30) | 782 (26) | 579 (31) | 159 (17) |  | 523 (21) |  | 1,305 (24) | 528 (33) |  |
|  | Unknown | 527 | 289 | 187 |  | 479 |  | 1,006 | 1,701 |  |

Key: HPV, human papilloma virus; OPC, oropharyngeal cancer; OC, oral cancer.
\* Never/Former category used when participants were only asked if they were a current drinker (HN5000 study). For these participants there was no information available to determine whether they were a former or a never drinker if they were not a current drinker.

HPV16 E6 serology was prioritized as a marker of HPV(+) oropharyngeal cancer as this has been shown to be a highly sensitive and specific marker of HPV oncogenic infection in oropharyngeal cancer and can be easily assayed from blood [38, 39]. Multiplex serology was performed on 75% (n= 5,294) of all HNC cases and 92% (n= 3,250) of oropharyngeal cancer cases using a previously developed Luminex assay [40, 41]. Multiplex serology generates quantitative data expressed in median fluorescence intensity (MFI) units for each pathogen-specific antigen and serum. Seropositivity for every antigen was based on previously determined standardized cut-offs in order to optimize sensitivity and specificity [42, 43].

When serology was missing, two concordant tumor markers, p16 immunohistochemistry (IHC) and high-risk HPV DNA in-situ hybridization (ISH) were required to determine HPV status. If p16 IHC and HPV DNA ISH were discordant or only one marker was available, then HPV status was classified as unknown. This algorithm was based on evaluation of biomarkers performance compared to molecular reference method (serology) from known data, led by consortium members [39, 44].

All samples (6,034 cases and 6,585 controls) were genotyped as part of the oral and pharynx cancer OncoArray study, except for 1,023 controls from the Toronto study which were genotyped as part of the Lung OncoArray. Tumor DNA sequencing data are available for 999 patients for whom adequate tumor tissue samples were present. Further details on the statistical analyses, genotyping and tumor sequencing methods are available in the Supplementary Methods (**Appendix A**).

## Results

### VOYAGER Data Resource: Clinical and Demographic Profile

The VOYAGER consortium comprises 10,530 participants in total (7,233 cases and 3,297 controls). The primary focus of the consortium was on oral cavity (N=2,975) and oropharyngeal (N=3,514) cancers, but a smaller number of cases from other head and neck subsites (N=744) were also included. The majority of cancer cases were contributed by HN5000 (41%), followed by Toronto (23%), Pittsburgh (13%), CHANCE (12%) and ARCAGE (11%). HPV status is available for 78% of all cases, tumor sequencing data for 14%, and the majority of cases was diagnosed with stage III or IV disease (**Appendix B, Table B.1**). The controls included were used for the genotyping studies noted above. To facilitate the use of this resource, we present here a brief description of the oral cavity and oropharyngeal cancers and controls, while noting that this resource contains additional head and neck subsites, described in **Appendix B, Table B.1,** that may be valuable to the scientific community.

The included controls differ significantly from the oral cavity and oropharyngeal cancer cases in all evaluated demographic variables and established risk factors, except for age (p= 0.308) (**Tables 1; Table 2**). HPV16 E6 serology status is available for 3,250 (92%) oropharyngeal cancer cases, of which 61% are HPV(+) (**Figure 1**; **Table 1**). When serology was missing, p16 and high-risk HPV DNA ISH concordance determined HPV oropharyngeal cancer status in the remaining ∼8% of cases. There was good concordance between p16 and HPV DNA ISH.

There are in total 2,975 oral cavity and 3,514 oropharyngeal cancer cases (2,138 HPV(+), 1,146 HPV(-) and 230 HPV status unknown oropharyngeal cancer cases) included in VOYAGER (**Table 1**). Most cases presented in males (73%), outnumbering females across all subsites. The overall mean age at diagnosis in all cases is 60 years (SD= 10.7), with the lowest mean age observed in the HPV(+) oropharyngeal cancer group (58 years (SD= 8.9)). Overall age at diagnosis for oropharyngeal cancer (59 (SD= 9.6)) is significantly younger than that of oral cavity (61 (SD= 12.2)) (p< 0.0001) (**Table 1**). Only 3% of oral and oropharyngeal squamous cell carcinoma cases (n= 224) presented under 40 years old, and these are predominantly patients with cancer of the oral cavity (71%). The highest frequency of postsecondary education is observed in the HPV(+) oropharyngeal cancer group (42%), with the lowest in the HPV(-) oropharyngeal group (28%) (**Table 1**). In general, the HPV(+) and HPV(-) oropharyngeal cancer cases differ significantly from each other in almost all evaluated demographic, clinical and risk factor variables (**Table 1**; **Table 2**).

Most cases are current (33%) or former smokers (42%). However, significant differences in smoking status are seen between the subsites (p< 0.0001) with the HPV(+) oropharyngeal group including the largest frequency of never smokers (34%) (**Table 2**). Similarly, more than half (63%) of the cases reported current alcohol drinking, but there are significant differences in alcohol drinking status between the subsites. Being overweight is most common among the HPV(+) oropharyngeal cancer cases (40%), with underweight being the least frequently recorded category across the oral and oropharyngeal cancer cases.

Most of the included cases presented late, at stage IV (57%), with 72% of oropharyngeal cancer presenting at stage IV versus only 39% of the oral cavity cancer cases (**Table 1**). Non-surgical treatment using radiotherapy, with or without chemotherapy, is the most common treatment modality in the oropharyngeal cancer group (66%), while surgery is the most common treatment modality in the oral cavity cancer group, with almost half (47%) of these patients receiving surgery alone and another 40% undergoing surgery plus adjuvant radiotherapy with or without chemotherapy (**Table 1**).

Collecting information on disease outcome was a primary focus of the consortium given the high risk of recurrence and poor survival associated with HNC. The median length of follow-up time for the cases is 5.3 years. The median overall survival time is 9.8 years and the five-year overall survival rate is 66% (**Table 3**). Patients with HPV(+) oropharyngeal cancer show the best overall survival, with a median survival time of 14.3 years and 5-year survival rate of 81% (**Table 3** and **Figure 2**). For oropharyngeal cancer patients with unknown HPV status, the probability of being alive at 5 years falls between that of HPV(+) and HPV(-) oropharyngeal cancer patients, at 63%, indicating these are likely a mix of HPV(+) and HPV(-) oropharyngeal cancer patients. The probability of being progression-free at 5 years after diagnosis is highest for the HPV(+) oropharyngeal cancer patients (75%) while it ranges from 44% – 50% for the other subsites. Disease-specific survival shows similar trends across subsites (**Table 3** and **Figure 2**). The HPV(+) oropharyngeal cancer patients have the highest probability of disease-specific survival at 5 years (88%) and those with HPV(-) oropharyngeal cancer the lowest (67%). Across all cancers, the probability of disease-specific survival was 78% (**Table 3** and **Figure 2**).

**Figure 2.**
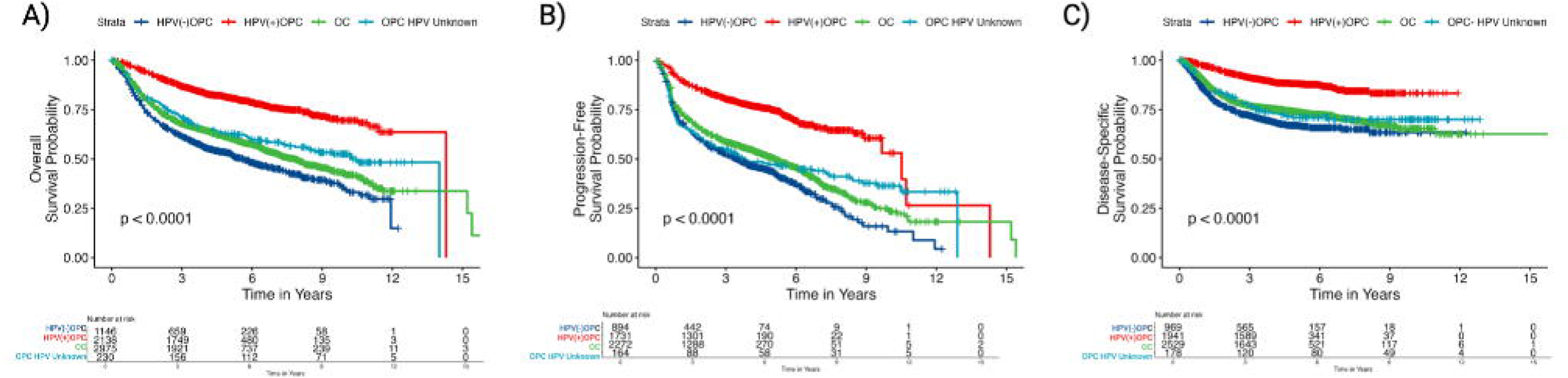
a) Overall survival, b) progression-free survival, and c) disease-specific survival for oral and oropharyngeal cancer cases in VOYAGER.

**Table 3.** Survival outcomes for the oral and oropharyngeal cancer cases in VOYAGER, stratified by subsite and HPV status.

|  | OPC HPV Unknown | HPV(+) OPC | HPV(-) OPC | OC | Total OPC + OC |
| --- | --- | --- | --- | --- | --- |
| Overall survival |  |  |  |  |  |
| N (number of cases) | 230 | 2,138 | 1,146 | 2,975 | 6,489 |
| Median overall survival time, in yrs | 10.3 years | 14.3 years | 5.6 years | 7.8 years | 9.8 years |
| Probability of survival at 5y (95%CI), % | 62.8 (56.7, 69.5) | 81.1 (79.4, 82.9) | 53.1 (50.2, 56.2) | 60.7 (58.9, 62.5) | 66.1 (65.0, 67.4) |
| Log-rank p-value | <0.001 |  |  |  |  |
| Progression-free survival |  |  |  |  |  |
| N (number of cases) | 164 | 1,731 | 894 | 2,272 | 5,061 |
| Median progression-free survival time, in yrs | 3.8 years | 10.5 years | 3.5 years | 5.1 years | 6.3 years |
| Probability of progression-free survival at 5y (95%CI), % | 47.2 (40.1, 55.7) | 74.9 (72.7, 77.1) | 43.7 (40.2, 47.3) | 50.5 (48.3, 52.7) | 57.4 (56.0, 58.9) |
| Log-rank p-value | <0.001 |  |  |  |  |
| Disease-specific survival* |  |  |  |  |  |
| N (number of cases) | 220 | 2,112 | 1,102 | 2,877 | 6,311 |
| Disease-specific survival at 5y (95%CI), % | 72.7 (66.8, 79.2) | 88.4 (87.0, 89.8) | 67.5 (64.6, 70.6) | 74.6 (72.9, 76.3) | 78.1 (77.0, 79.2) |
| Gray's p-value | <0.001 |  |  |  |  |

Key: HPV, human papilloma virus; OPC, oropharyngeal cancer; OC, oral cancer; Overall, including oral and oropharyngeal cancers.
Progression-free survival data was not available for the CHANCE study.
\*Median not reached for disease-specific survival.
HPV16 E6 serology was prioritized as a marker of HPV(+) oropharyngeal cancer. When serology was missing, two concordant tumor markers, p16 immunohistochemistry (IHC) and high-risk HPV DNA in-situ hybridization (ISH) were required to determine HPV status.

Median length of follow-up time differs between the five studies, but not significantly so (**Appendix B Table B.1;** p= 0.400). Patients from the ARCAGE study have lower overall, progression-free and disease-specific survival compared to the other studies for HPV(+) oropharyngeal cancers, however, there are only 63 HPV(+) oropharyngeal cancer cases in ARCAGE (**Appendix C Figures C.1 - 3**). In addition, smoking rates in ARCAGE are high compared to other studies and this has been shown to affect interpretation of its HPV(+) oropharyngeal cancer profiles [38].

The clinical, demographic and survival features of the VOYAGER cohort are characteristic of what we know of HNC patients, epidemiologically and clinically [4, 45, 46]. We show the predominance of male patients, the established differences in age at diagnosis, education and survival of HPV driven versus non-HPV cancers, and the expected distributions of risk factor exposures [4, 45]. Therefore, VOYAGER provides a reliable representative resource for further research into this patient population. Importantly, the combination of clinical follow-up information, somatic tumor sequencing and germline genotyping is a particular strength of this resource (**Figure 3**).

**Figure 3.**
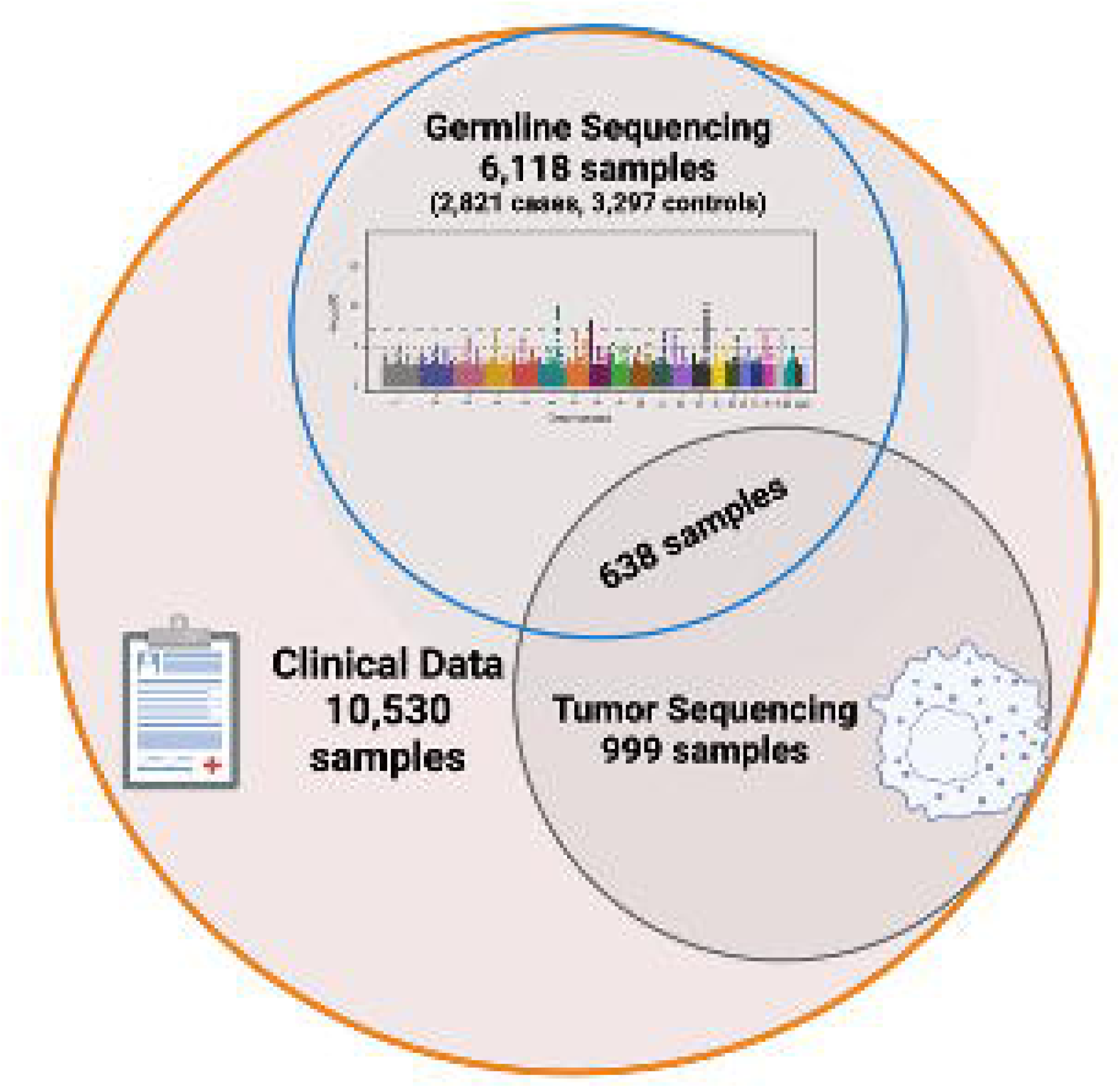
Overlapping data available in VOYAGER. Genotyping data includes cases and controls, tumor sequencing samples are for oropharyngeal and oral cavity cases only, and clinical data includes all available cases and controls. *Created in BioRender. (2026)* https://BioRender.com/h48b673

## Discussion

Several important findings have already come from the VOYAGER consortium. Previous GWAS studies of oral and oropharyngeal cancer conducted by our group have highlighted the important role of the human leukocyte antigen (HLA) region (6p21.3) in susceptibility to oropharyngeal cancer [47]. Specifically, a two-fold protective effect was observed for oropharyngeal cancer and the HLA haplotype DRB1*1301-DQA1*0103-DQB1*0603. This haplotype was previously reported to also be protective for cervical cancer, a cancer type that is primarily driven by HPV infection [47]. In VOYAGER, HPV status was determined for 92% of the oropharyngeal tumors via HPV16 E6 serology enabling the first GWAS focused on HPV driven HNCs using fine mapping techniques [48]. Within the 6p21.3 locus, there were two specific loci (rs4713462 and rs9269942) independently associated with reduced risk of oropharyngeal cancer (**Appendix C Figure C.4**). These loci were separately associated with antibodies against specific HPV16 proteins which implicates specific germline variants in the natural immune response against HPV(+) oropharyngeal cancer, supporting the use of therapeutic vaccines to protect against HPV infections and help prevent this disease.

In addition, a recent publication utilized VOYAGER data to develop a risk prediction model for HNC including genetic markers, HPV serostatus, demographic and lifestyle risk factors in populations of European ancestry. The addition of HPV serology provided substantial predictive accuracy for oropharyngeal cancer (AUC= 0.94, 95%CI: 0.92 – 0.95 in men and AUC= 0.92, 95%CI: 0.88 – 0.95 in women) above that of previously published models, highlighting the need to consider primary prevention and intensive surveillance for oropharyngeal cancer subgroups [49]. Importantly, however, while HPV serology is a marker for HPV driven oropharyngeal cancer, the use of HPV serology needs to be carefully evaluated among smokers as demonstrated in another VOYAGER publication [38].

Diagnostic accuracy of HPV serology was evaluated utilizing VOYAGER data and found to be highly sensitive and specific independent of age, sex, year of diagnosis, BMI at diagnosis, current alcohol use and primary tumor size, but exhibited some variation in diagnostic accuracy for heavy smokers and by lymph node involvement. This work provides further evidence that this additional HPV biomarker can be used for early diagnosis.

Randomized clinical trials remain the ‘gold standard’ to ascertain the causal effect of interventions or modifiable exposures. However, they are not always feasible in terms of cost, time or ethics [50]. Conversely, observational studies are subject to confounding, bias and reverse causality. This has significantly limited study design in HNC research, given that it is a relatively rare cancer with correlated risk factors, requiring long-term follow-up from the point of exposure to disease onset. To overcome such limitations, Mendelian randomization (MR) uses measured genetic variation to examine the causal effect of potentially modifiable exposures on health outcomes in observational data [51–53]. The genotyping data available in VOYAGER has contributed to multiple MR studies [54–59] investigating a wide range of genetically proxied exposures. These studies have among other things demonstrated an independent causal effect for both smoking and alcohol on both HPV(+) and HPV(-) HNC, suggesting that the effect of alcohol consumption may have been previously underestimated [54, 58]. This work further strengthens the evidence to support public health messaging around prevention in HNC.

## Conclusion and future prospects

The value of HNC data generated as part of VOYAGER highlights its use for studies focused on prognosis, particularly overall survival where data is most complete. The variation across centers is an important consideration, as studies contributing to VOYAGER were conducted across different geographical settings and health systems. The VOYAGER data resource is particularly suited for genomic studies on risk factors and outcome.

## Author contributions

This manuscript was conceived by M.G., S.V. and B.D. Formal analyses and visualization of results for this manuscript were conducted by M.G. and S.V. The manuscript was drafted by M.G., S.V. and A.A. All authors contributed to the interpretation of the review and editing of the manuscript. All authors read and approved the final manuscript.

## Data Availability

For VOYAGER data access requests, please contact. Non-commercial research projects are generally approved if the proposed research complies with the signed agreements between the studies and their research participants.
Genotype data for the oral and pharynx cancer OncoArray study have been deposited at the database of Genotypes and Phenotypes (dbGaP) and are available under controlled access under accession phs001202.v1.p1. Genotype data for the All of Us study are also available via dbGaP under controlled access under accession phs003225.v1.p1.
The oral and pharyngeal GWAS summary statistics by cancer site and world region have been deposited in the IEU Open GWAS platform (https://gwas.mrcieu.ac.uk/) under the GWAS IDs: ieu-b-89, ieu-b-90, ieu-b-94, ieu-b-96, ieu-b-93, ieu-b-97, ieu-b-91, ieu-b-95 and ieu-b-98.

https://voyager.iarc.who.int/

https://www.ncbi.nlm.nih.gov/projects/gap/cgi-bin/study.cgi?study_id=phs001202.v1.p1

https://www.ncbi.nlm.nih.gov/projects/gap/cgi-bin/study.cgi?study_id=phs003225.v1.p1

## Acknowledgements

The authors would like to thank all study participants.

## List of abbreviations

ARCAGE: Alcohol-related cancers and genetic susceptibility in Europe
AJCC: American Joint Committee on Cancer
ABRA: Assembly-based ReAligner
BWA: Burrow-Wheeler Aligner
CHANCE: Carolina Head and Neck Cancer Epidemiology
GAME-ON: Genetic Associations and Mechanisms in Oncology
GWAS: Network Genome-wide association study
HNC: Head and neck cancer
HN5000: Head and Neck 5000 study
HPV: Human papillomavirus
IHC: Immunohistochemistry
ISH: In situ hybridization
MFI: Median fluorescence intensity
MSH-PMH: Mount Sinai Hospital-Princess Margaret
NGS: Next Generation Sequencing
OC: Oral cancer
OPC: Oropharyngeal cancer
UNMASC: Unmatched Normals and Mutant Allele Status Characterization
VOYAGER: human papillomaVirus, Oral and oropharYngeal cAncer GEnomic Research

## Ethics approval and consent to participate

The VOYAGER project was approved by the International Association for Research on Cancer (IARC) Ethics Committee (IEC) (Project No.: 16 – 34). The five parent studies were all approved by the respective institutional review boards (IRBs) at each of the participating centers: University of North Carolina at Chapel Hill (Study No.: 01-0390), University of Pittsburgh (STUDY19120160), The University Health Network (Project No.: 07-0521), Sinai Health System (REB No.: 08-0191) and the National Health Service, Health Research Authority (Project ID: 24028).

## Consent for publication

Not applicable.

## Availability of data and materials

For VOYAGER data access requests, please contact. Non-commercial research projects are generally approved if the proposed research complies with the signed agreements between the studies and their research participants.

Genotype data for the oral and pharynx cancer OncoArray study have been deposited at the database of Genotypes and Phenotypes (dbGaP) and are available under controlled access under accession phs001202.v1.p1. Genotype data for the All of Us study are also available via dbGaP under controlled access under accession phs003225.v1.p1.

The oral and pharyngeal GWAS summary statistics by cancer site and world region have been deposited in the IEU Open GWAS platform (https://gwas.mrcieu.ac.uk/) under the GWAS IDs: ieu-b-89, ieu-b-90, ieu-b-94, ieu-b-96, ieu-b-93, ieu-b-97, ieu-b-91, ieu-b-95 and ieu-b-98.

