## Appendices for "VOYAGER: an international consortium investigating the role of human papilloma virus and genetics in oral and oropharyngeal cancer risk and survival"

**Appendix A: Supplementary Methods**

**Statistical description of the VOYAGER Data Resource**

Continuous variables were described using mean and standard deviation. Categorical variables were described as counts and percentages (%). Independent two-sample t-tests were used to evaluate differences between continuous variables and Chi-square tests for categorical variables using the *ttest* and *tabi* function in Stata. Descriptive analysis was performed using Stata Statistical Software (Release 18, College Station, TX: StataCorp LLC). Median follow-up time was estimated using the product-limit estimator, calculated potential follow-up time distribution based on the Kaplan-Meier method applied to the censored times, reversing the roles of event status and censored. Probability of overall and progression-free survival was evaluated using Kaplan-Meier analysis, and differences between groups were tested with a log-rank test. Progression-free survival was evaluated with progression defined as relapse, recurrence, metastasis or death events. Cancer-specific survival was evaluated using Fine-gray models for competing risks of cancer-specific deaths versus all other causes to evaluate cancer-specific risk of mortality. Survival analysis was conducted using R version 4.3.2 using the *survival* and *tidycmprsk* packages [1, 2].

**Genotyping and tumor sequencing methods**

Genotyping data was generated at the Center for Inherited Disease Research (CIDR) in several rounds. The first round (X01HG007780) was performed using the Illumina OncoArray, which was custom designed for cancer studies by the OncoArray Consortium part of the Genetic Associations and Mechanisms in Oncology (GAME-ON) Network. All samples (6,034 cases and 6,585 controls) were genotyped as part of the oral and pharynx cancer OncoArray study, except for 1,023 controls from the Toronto study which were genotyped as part of the Lung OncoArray. This genotyping data was used to conduct the first genome-wide association study (GWAS) on head and neck cancer in 2017 [3]. With the generation of HPV status information based on serology, these genotyping data were also used to run a GWAS of oral and oropharyngeal cancer, stratified by HPV status [4]. A second round of genotyping was undertaken (X01HG010743) for an additional 1,395 samples in VOYAGER. This was conducted on the All of Us Array, an Illumina array customized for the All of Us Consortium and designed to include multiethnic context [5]. The genotyping data from OncoArray and the All of Us Array, recently contributed to the largest HNC GWAS to date including 19,073 cases and 38,357 controls identifying 29 independent genetic loci [6].

Tumor DNA sequencing data are available for 999 patients in the VOYAGER consortium for whom adequate tumor tissue samples were present. These data were generated using a custom cancer gene panel that has been previously reported [7-11]. Next generation sequencing (NGS) was performed using the Agilent SureSelect protocol and reagents according to manufacturer’s specifications. The assay targets all genes of the Cancer Gene Census, in addition to clinically relevant targets such as drug metabolizing enzymes. The assay also performs whole genome sequencing of HPV16 and 18 using methodology previously reported to offer clinical diagnostic accuracy comparable or better to conventional approaches while at the same time capturing base level resolution across the HPV genome [8]. The resulting libraries containing paired-end reads of 150 bases in length were sequenced on Illumina sequencers, primarily NovaSeq according to manufacturer’s specifications, with a mean target depth coverage of 500x. Further details of the assay can be found elsewhere [12]. All analytic tools and the pipelines for integrating steps have been publicly reported and are available as open source software including: Burrow-Wheeler Aligner (BWA) for sequence alignment [13], Next NGS Copy [12] for copy number assessment, Strelka [14], Assembly-based ReAligner (ABRA) for realignment and structural variant detection [15], and Unmatched Normals and Mutant Allele Status Characterization (UNMASC) for variant prioritization and filtering [7].

**Appendix B:**

**Table B.1: Description of the head and neck cancer cases included in the VOYAGER consortium, overall and stratified by study**

|  | Total N  (%)* | ARCAGE  N (%)** | CHANCE  N (%)** | HN5000  N (%)** | Pittsburgh  N (%)** | Toronto  N (%)** |
| --- | --- | --- | --- | --- | --- | --- |
| N cases | **7,233** | 826 (11) | 862 (12) | 2,992 (41) | 910 (13) | 1,643 (23) |
| HPV serology | **5,472 (76)** | 675 (12) | 509 (9) | 2,952 (54) | 372 (7) | 964 (18) |
| HPV p16 | **3,575 (49)** | 424 (12) | 458 (13) | 1,178 (33) | 442 (12) | 1,073 (30) |
| HPV DNA ISH | **595 (8)** | 0 (0) | 0 (0) | 200 (34) | 395 (66) | 0 (0) |
| HPV final status*** | **5,641 (78)** | 675 (12) | 509 (9) | 2953 (52) | 540 (10) | 964 (17) |
| Tumor DNA sequencing | **999 (14)** | 109 (11) | 124 (13) | 393 (39) | 133 (13) | 240 (24) |
| Genotype – OncoArray | **4,353 (60)** | 652 (15) | 724 (16) | 1,084 (25) | 809 (19) | 1,084 (25) |
| Genotype – All of Us | **1,395 (19)** | 71 (5) | 0 (0) | 0 (0) | 339 (24) | 985 (71) |
| Anatomical site |  |  |  |  |  |  |
| Oropharynx | **3,514 (49)** | 278 (8) | 346 (10) | 1,598 (45) | 389 (11) | 903 (26) |
| Oral cavity | **2,975 (41)** | 419 (14) | 348 (11) | 1,148 (39) | 493 (17) | 567 (19) |
| Hypopharynx | **357 (5)** | 70 (20) | 30 (8) | 185 (52) | 17 (5) | 55 (15) |
| Larynx | **107 (1)** | 0 (0) | 92 (86) | 8 (7) | 1 (<1) | 6 (6) |
| Unknown primary | **103 (1)** | 0 (0) | 0 (0) | 16 (16) | 0 (0) | 87 (84) |
| Pharynx, nos | **72 (<1)** | 13 (18) | 46 (64) | 7 (10) | 5 (7) | 1 (1) |
| Overlapping | **44 (<1)** | 43 (98) | 0 (0) | 0 (0) | 0 (0) | 1 (2) |
| External lip | **26 (<1)** | 0 (0) | 0 (0) | 23 (88) | 0 (0) | 3 (12) |
| Other site**** | **35 (<1)** | 3 (9) | 0 (0) | 7 (20) | 5 (14) | 20 (57) |
| Stage (AJCC 7^th^ ed.) |  |  |  |  |  |  |
| In situ | **4 (<1)** | 0 (0) | 0 (0) | 0 (0) | 1 (25) | 3 (75) |
| I | **1037 (14)** | 102 (10) | 146 (14) | 488 (47) | 154 (15) | 147 (14) |
| II | **1010 (14)** | 127 (13) | 144 (14) | 451 (45) | 139 (14) | 149 (14) |
| III | **936 (13)** | 124 (13) | 147 (16) | 352 (38) | 130 (14) | 183 (19) |
| IV | **4025 (56)** | 293 (7) | 425 (11) | 1,693 (42) | 467 (12) | 1,147 (28) |
| Unknown | **221 (3)** | 180 (81) | 0 (0) | 8 (4) | 19 (9) | 14 (6) |
| Median follow up time (years) | **5.4** | 8.4 | 10.1 | 4.9 | 5.7 | 5.2 |
| * % of total N in VOYAGER with that information (column).  ** % of total N with that information (row).  *** HPV16 E6 serology was prioritized as a marker of HPV(+) oropharyngeal cancer. When serology was missing, two concordant tumor markers, p16 immunohistochemistry (IHC) and high-risk HPV DNA in-situ hybridization (ISH) were required to determine HPV final status.  **** Other site: nasal cavity n=1, nasopharynx n= 4, other ear/ bone/ skin/ lacrimal duct and gland n= 4, overlapping lesion n= 3, paranasal sinuses n= 4, salivary glands n= 2, head and neck, not otherwise specified (nos) n= 13, carcinoma in situ n= 4. | | | | | | |

**Appendix C: Supplementary Figures**

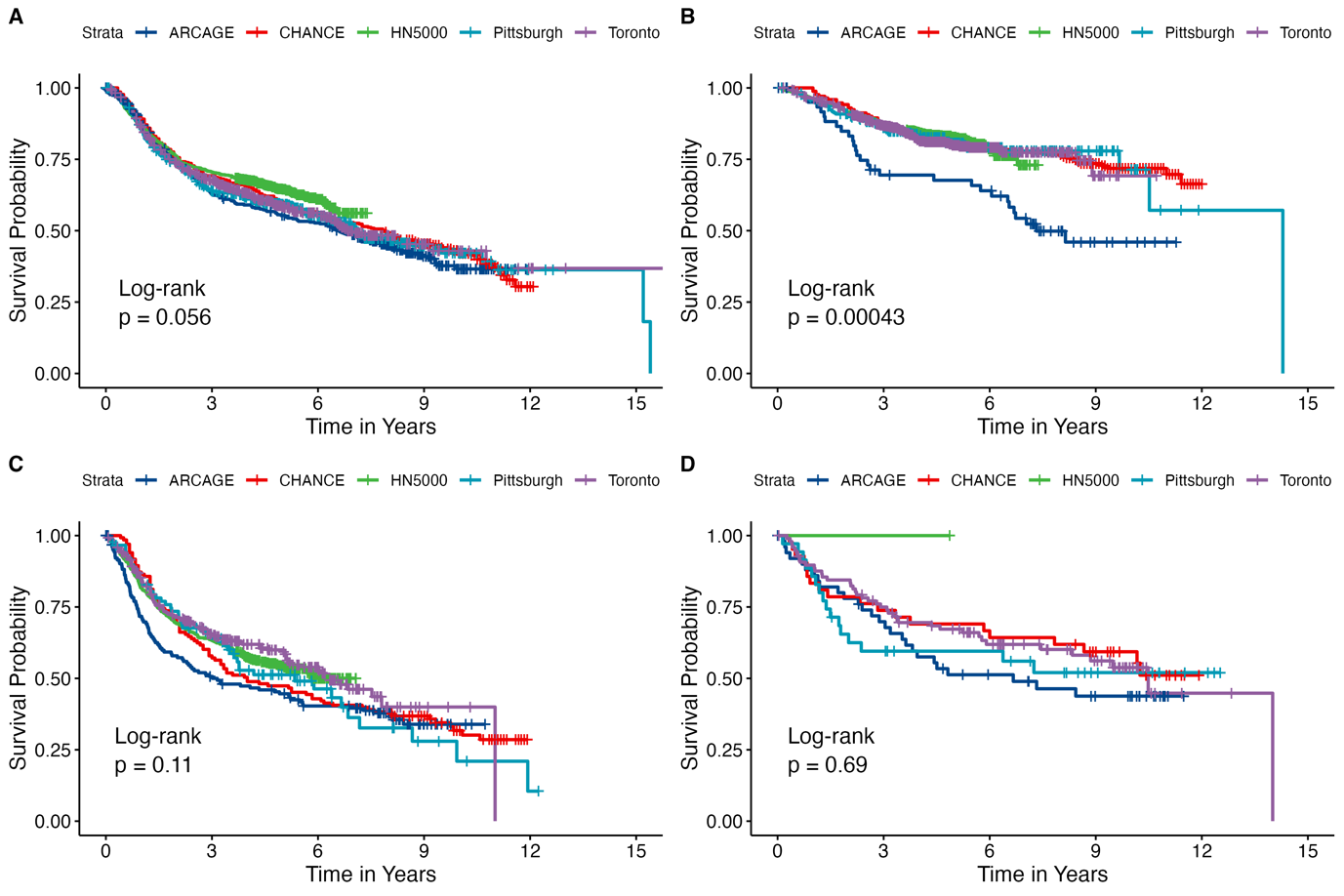

**Figure C.1:** Overall survival by study for A) oral cavity; B) HPV(+) oropharyngeal cancer; C) HPV(-) oropharyngeal cancer; D) oropharyngeal cancer with HPV status unknown

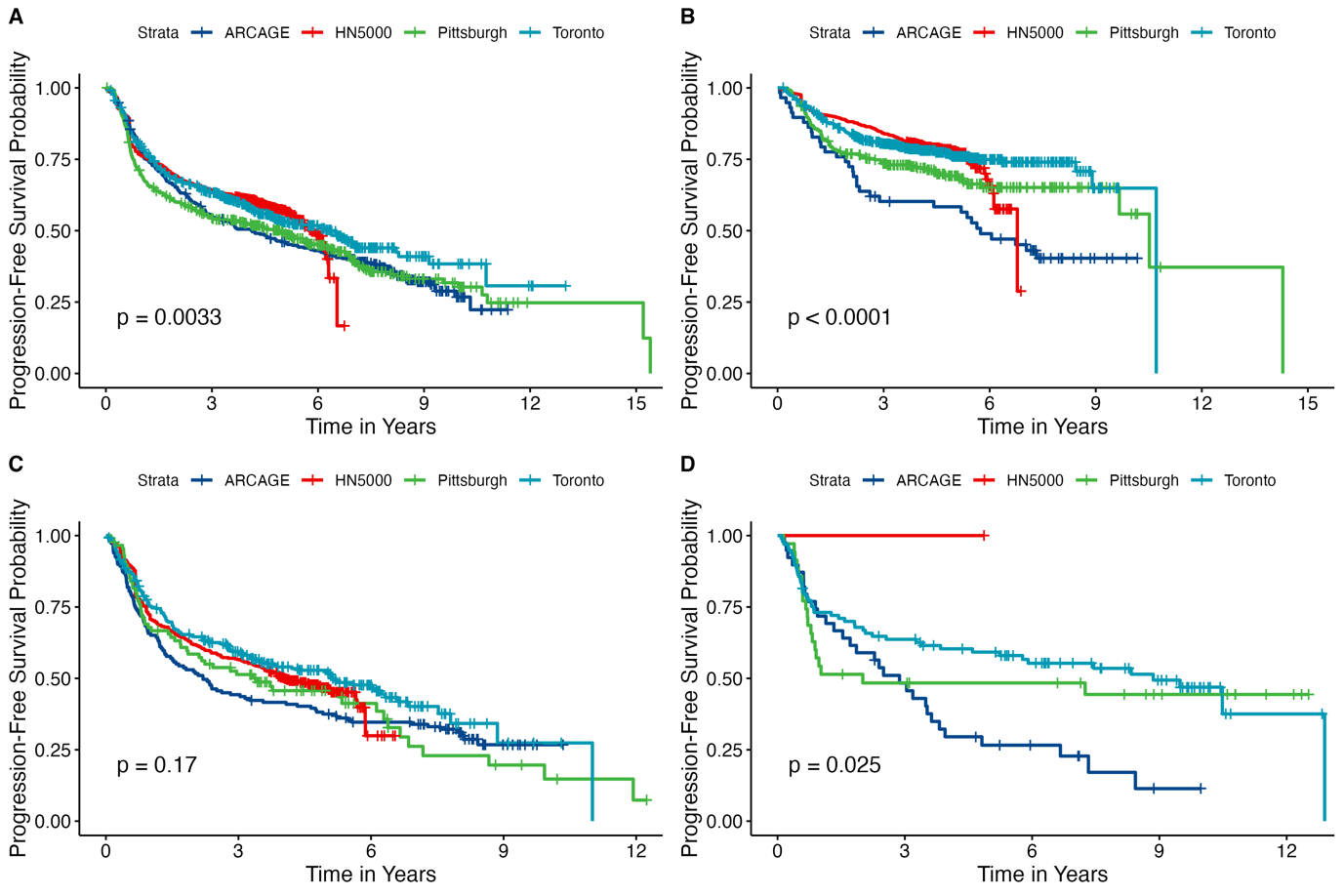

**Figure C.2:** Progression-free survival by study for A) oral cavity; B) HPV(+) oropharyngeal cancer; C) HPV(-) oropharyngeal cancer; D) oropharyngeal cancer with HPV status unknown. Progression events other than death were not available for the CHANCE study.

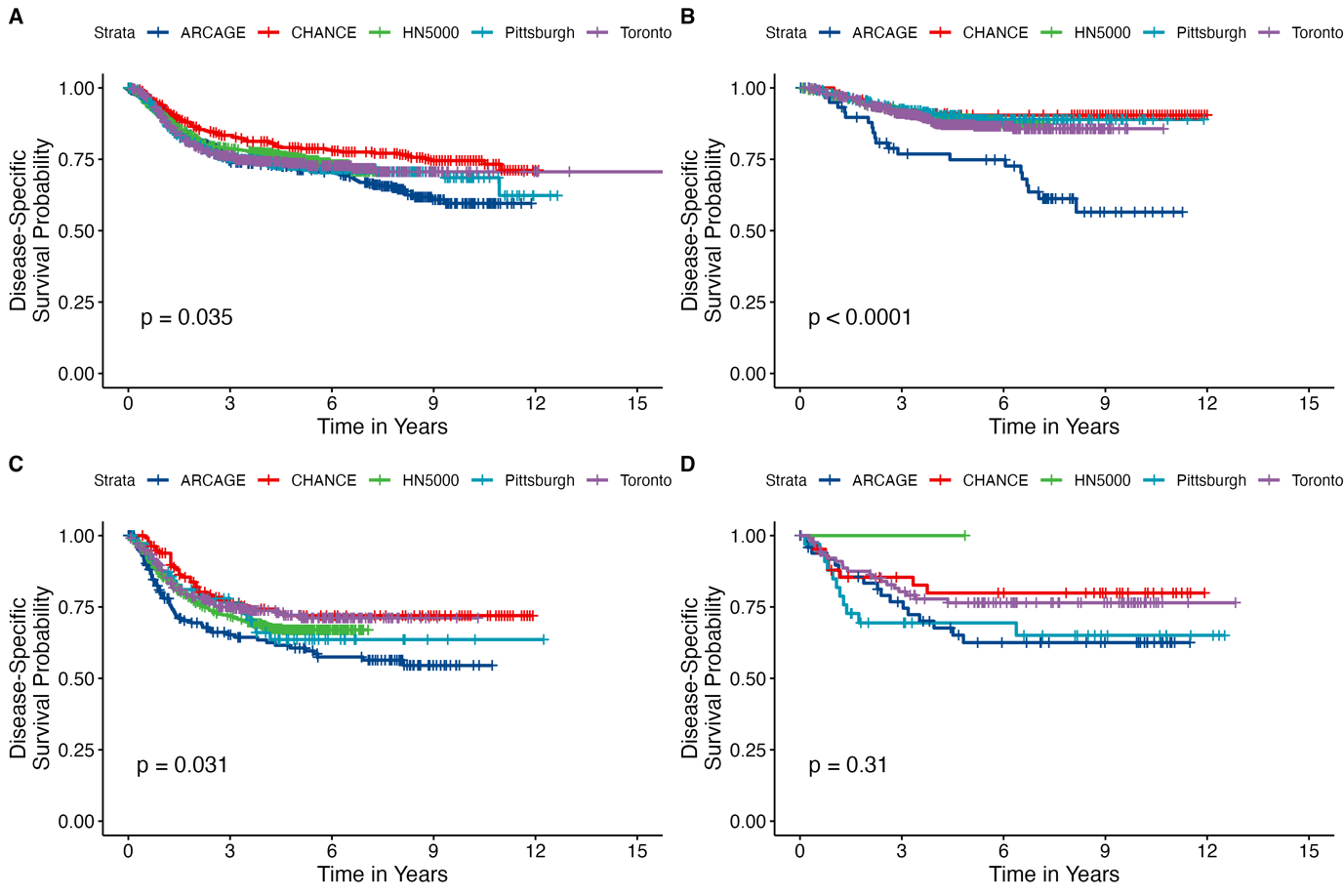

**Figure C.3:** Cancer specific survival by study for A) oral cavity; B) HPV(+) oropharyngeal cancer; C) HPV(-) oropharyngeal cancer; D) oropharyngeal cancer with HPV status unknown

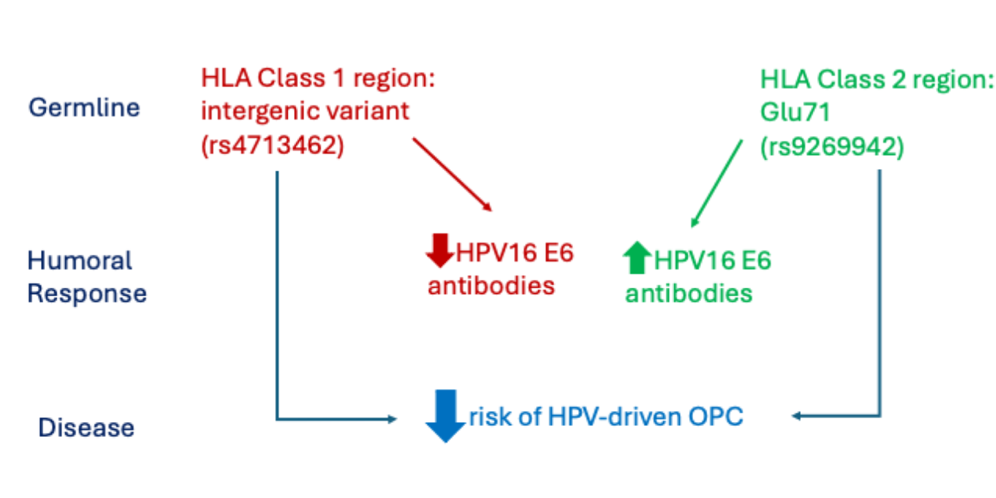

**Figure C.4:** Viral host interactions suggest HLA loci that are specific for HPV16 viral proteins*.*

**Appendix D: Supplementary References**

1.	Sjoberg DD. tidycmprsk: Competing Risks Estimation. R package version 1.1.0, 2024 [Available from: <https://github.com/MSKCC-Epi-Bio/tidycmprsk>, <https://mskcc-epi-bio.github.io/tidycmprsk/>.

2.	Therneau T. A Package for Survival Analysis in R. R package version 3.8-3,.

<https://CRAN.R-project.org/package=survival>
